# Development of ADAF screening tool for emotional and coping problems in cancer patients

**DOI:** 10.1101/2021.01.26.21250516

**Authors:** Pedro Pérez-Segura, Santos Enrech Francés, Ignacio Juez Martel, Maria Angeles Pérez Escutia, Elena Hernández Agudo, Leticia Leon Mateos, Guido Corradi, Helena Olivera Pérez-Frade, Francisco Sánchez Escamilla, Marta Baselga López, Jose Luis Baquero, Marta Redondo Delgado

## Abstract

**Purpose:** Psychological screening in patient with cancer is recommended by clinical guidelines, however most of scales have large number of items, difficulty detection and refer from routine consultations. The specific objective of the study was to develop and validate the ADAF screening for anxiety, depression and coping.

**Methods/Patients:** Cross-sectional, multicenter study performed in the medical and radiotherapy oncology services of 5 hospitals in Madrid, coordinated by the Medical Oncology Service of the Hospital Clínico San Carlos (CEIC nº19 / 265-E). To determine the psychometric properties, the ADAF screening questionnaire ADAF was administered, including 5 items (one related to anxiety symptoms, two related to depressive symptoms, one for helplessness coping and one for avoidance coping), and as a gold-standard the HADS and the MiniMAC. Intraclass correlation coefficients and receiver operating characteristic curves were performed. The p value <0.05 was considered significant.

**Results:** A total of 186 patients completed the evaluation. The correlation coefficients were significant for all dimensions (Anxiety, Depression, Helplessness coping, and Avoidance Coping), with p <0.001. The statistical analysis of ROC curves suggests that the cut-off point for screening is equivalent to a score > 2 points (3 in the case of depression, having two items), with a sensitivity and specificity between 62 and 90%, depending on the item, and an area under the curve above 0.8 for the first 4 items.

**Conclusions:** ADAF screening has adequate reliability, good sensitivity and specificity. This instrument is useful and easy to use to identify emotional and coping problems in cancer patients.

## Introduction

Patients with cancer often experience negative emotions such as sadness, anxiety, and fear which, if unaddressed, can impair adaptation and emotional well-being and may negatively affect treatment adherence.

In oncological patients, it is important to screen whenever possible for emotional problems, since 25%–40% will exhibit high levels of psychological distress requiring professional intervention [1,2]. A systematic review and meta-analysis shows the prevalence of major depression (15%), minor depression (20%), and anxiety disorders (10%) in patients treated for cancer [2]. Therefore, distress has been endorsed as the “6th vital sign” in cancer care and recommendations of routine screening for management of distress have been established as an integral part of whole-person cancer care in clinical practice guidelines [3,4]. In Oncology, emotional distress (in its main components of anxiety and depression) is expected during the entire disease trajectory. It is a source of suffering on its own, but it may also interfere compliance with treatment, as well as with both health and well-being.

The well-documented high level of psychological variables morbidity among cancer patients has fueled interest in mechanisms of coping [5]. Coping refers to the set of cognitive and behavioural responses implemented after a stressful event in an attempt to mitigate its psychological impact. The different ways of coping have been grouped into large strategies. Of all of them, those that have been most studied in cancer, due to its effect on psychological well-being and on the disease evolution, have been passive (or helpless) and avoidant coping. [6–9]. If the diagnosis is interpreted as a loss, or defeat and death are seen as inevitable, a “helpless/hopeless” adjustment results. “Cognitive avoidance” appears when the threat is so great that people minimize, avoid or even deny its severity [10]. Studies have consistently found that patients with helpless/hopeless and anxious adjustment styles as cognitive avoidance have greater emotional distress [11-14].

Patients should be screened for emotional distress and maladaptive coping at their initial visit, at appropriate intervals and as clinically indicated, especially with changes in disease status (ie, post-treatment, recurrence, progression) and when there is a transition to palliative and end-of-life care, but there is a lack of instruments “easy-applied” in the routine ambulatory setting to asses this psychological needs. In a study in health professionals working in cancer care, they suggested that ideal screening practice was to use one, two or three simple questions or a short validated questionnaire, but not to refer to a specialist for a diagnosis. The main barrier to successful screening was lack of time, but insufficient training and low confidence were also influential [15].

The aim of this study was to assess the reliability and validity of the ADAF screening in outpatient care through patients with cancer.

## Methods

### Study design and patients

A validation cross-sectional study was conducted. This study was carried out in the medical and radiotherapy oncology services of 5 hospitals of the public health system of the Community of Madrid. The study was coordinated by the medical oncology service of the Hospital Clínico San Carlos.

The reference population consists of all individuals from these hospitals catchment areas. The inclusion criteria were patients with oncological disease, over 18 years of age, and having facilitated informed consent. Patients who did not met these criteria, presented cognitive impairment or were not able to carry on a conversation or understand the questions on the questionnaire were excluded. The research protocol was approved by the Ethics and Clinical Research Committee of the Hospital Clínico San Carlos (CEIC nº19 / 265-E).

After providing written informed consent, consecutive eligible patients were included during the scheduled visits to the medical or radiation oncology departments. Respondents filled out the questionnaires individually, but a health psychologist accompanied the patient in case of needing clarification. All collected data were registered anonymized in an electronic health record.

### Measures

The questionnaire requested descriptive data from each patient with sociodemographic variables including sex, age, nationality, marital status, educational level, employment status, and work activity.

The ADAF screening questionnaire was administered, along with the HADS and Mini-MAC. Intraclass correlation coefficients were calculated and ROC curve (receiver operating characteristic) analyses were performed. The value of p <0.05 was considered significant.

The ADAF screening questionnaire measures psychological problems in two dimensions: a) negative emotions, consists of three items (one related to anxiety symptoms, two related to depressive symptoms) and b) dysfunctional coping comprises the strategies that maintain or strengthen stressors. It consists of two items (one to deal with impotence and another to deal with avoidance). (Appendix 1) Items are scored on a four-point Likert scale from 0 (“almost never”) to 3 (“almost always”) based on a recall period of one week; higher scores represent poorer functioning.

It takes about 2 minutes to be completed by the patient or requested by the professional.

As Gold Standards we use the Mini-MAC scale and the HADS. The Mini-Mental Adjustment to Cancer scale (Mini-MAC) is among the most widely used instruments in assessing cancer-specific coping. The 29-item Mini-MAC [16], Spanish version [17,18] is a questionnaire derived from the original scale of Mental Adjustment to Cancer (MAC) [19]. This self-rated questionnaire examines five cancer-specific coping strategies: (1) Fighting spirit (the illness is experienced as a challenge and the patient has some degree of control over the situation (4 items); (2) Helplessness (the individual senses irreparable loss, fears death, and lacks insight into their situation (8 items); (3) Anxious preoccupation (the patient is afraid and doubts whether there is any possibility of exerting some control over the situation (8 items); (4) Cognitive avoidance (the threat and need for personal control are downplayed (4 items), and (5) Fatalism (the individual believes that their disease cannot be controlled and passively accepts it (5 items). Each item is scored using a 4-point Likert scale. The higher the subscale score, the more that coping strategy is used. The Spanish version of the scale was used for this study.

Negative emotions were assessed with the Hospital Anxiety and Depression Scale (HADS) [20]. Previous studies have shown the adequacy of HADS in evaluating emotional condition in the physical disease, and, specifically in cancer patients [21,22].

The HADS is a 14-item questionnaire with two dimensions (depression and anxiety), which has previously been validated as an appropriate instrument for use with Spanish cancer patients [21], showing its good psychometric characteristics and confirming its bifactorial structure, as proposed by Zigmond and Snaith [23].

### Statistical analysis

A description of the sociodemographic characteristics of patients included was explored with frequency distribution and the mean and standard deviation or median and percentiles.

The correlation between questionnaires sum scores (Pearson r) was calculated. We calculated operating characteristics comprising sensitivity, specificity, Youden index, positive predictive value, negative predictive value, overall accuracy (percentage of true results) for all possible cutoff points on the ADAF.

To assess the diagnostic accuracy of the HADS and MiniMAC and the ADAF, we performed ROC analyses. ROC curves indicate sensitivity and specificity combined for all possible cutoff points, such that the area under the curve (AUC) is a measure of diagnostic accuracy [24]. The AUC summarized the ability of the ADAF to discriminate between patients with and without negative emotions or dysfunctional coping. Higher AUC indicated better discriminatory capacity.

For all analyzes the level of statistical significance will be less than or equal to 0.05. Statistics were performed using the Jamovi and R statistical software 4.02.

## Results

A total of 195 patients consented to participate and completed the evaluation between July 2019 and October 2019. Baseline characteristics are summarized in Table 1. Females represented 73% (n = 122) of the sample with a mean age of 59 years [standard deviation (SD) = 13.1, range 18–85]; most were married or partnered (64.1%), and only a 33.3% had a primary level of education. The most common employment status was retired (51.3%), due to age or illness.

**Table 1.** Patient characteristics

| Variables | Value (n =195) |
| --- | --- |
| **Age, mean (S.D.), years** | 59 (13.1) |
| **Female, n (%)** | 122 (73) |
| **Marital status n (%)** |  |
| Single/Divorced | 52 (26.6) |
| Married/Partened | 125 (64.1) |
| Widow | 18 (9.2) |
| **Working status, n (%)** |  |
| Active | 40 (21) |
| Unemployed | 6 (3.1) |
| Student | 2 (1) |
| Retired | 100 (51.3) |
| Housewife | 19 (9.7) |
| Sick leave | 24 (12.3) |
| Not reported | 4 (2.1) |
| **Education n (%)** |  |
| Primary schooling | 66 (33.3) |
| Secondary | 59 (30.3) |
| University | 70 (36.4) |
| **HADS, mean (S.D.)** |  |
| Depression (0-21) | 4.60 (3.79) |
| Anxiety (0-21) | 7.29 (4.07) |
| **MiniMAC, mean (S.D.)** |  |
| Helplessness coping (8-38) | 12.9 (4.62) |
| Avoidance Coping (4-16) | 11.3 (3.03) |
| **ADAF, mean (S.D.)** |  |
| Depression (0-6) | 1.85 (1.73) |
| Anxiety (0-3) | 1.12 (0.96) |
| Helplessness coping (0-3) | 0.70 (0.89) |
| Avoidance Coping (0-3) | 1.51 (1.14) |

In our sample, screening with ADAF, a 14.5% showed positive for depression. Regarding anxiety, we found that a 32.8% presented anxiety. A total of 3.2% were classified as coping avoidance and 39.2% as coping helplessness.

Pearson correlations showed a statistically significant correlation between paired variables. Mini-MAC coping avoidance and ADAF coping avoidance showed the lowest correlation (r = .37, p < 0.001) and Mini-MAC coping helplessness and ADAF coping helplessness (r = .47, p < 0.001). While HADS depression and ADAF depression (r = .64, p < 0.001) and HADS anxiety and ADAF anxiety (r = .65, p < 0.001) showed the highest. For screening, ADAF had strong accuracy, with most areas under the curve (AUC) above 0.80 (anxiety AUC 0.85, depression AUC 0.84, coping Helplessness AUC 0.82), whereas performance was poor for identifying coping avoidance AUC of 0.68 (Figures 1-4).

**Figure 1.**
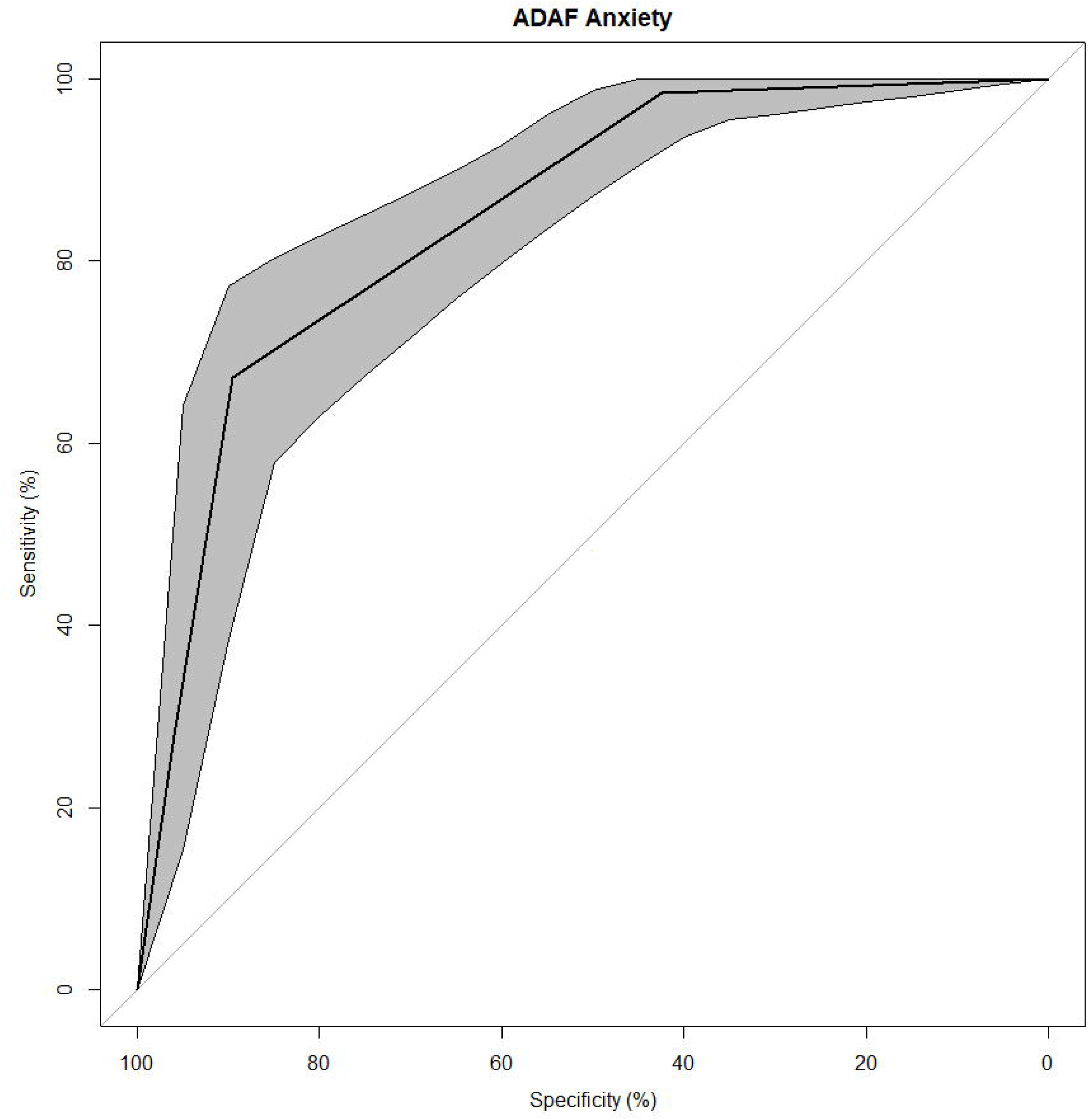
Receiver operating characteristic (ROC) curves for ADAF anxiety. Area under the ROC curve with 95% confidence interval (CI) is 0.85 (95% CI, 0.79-0.90).

**Figure 2.**
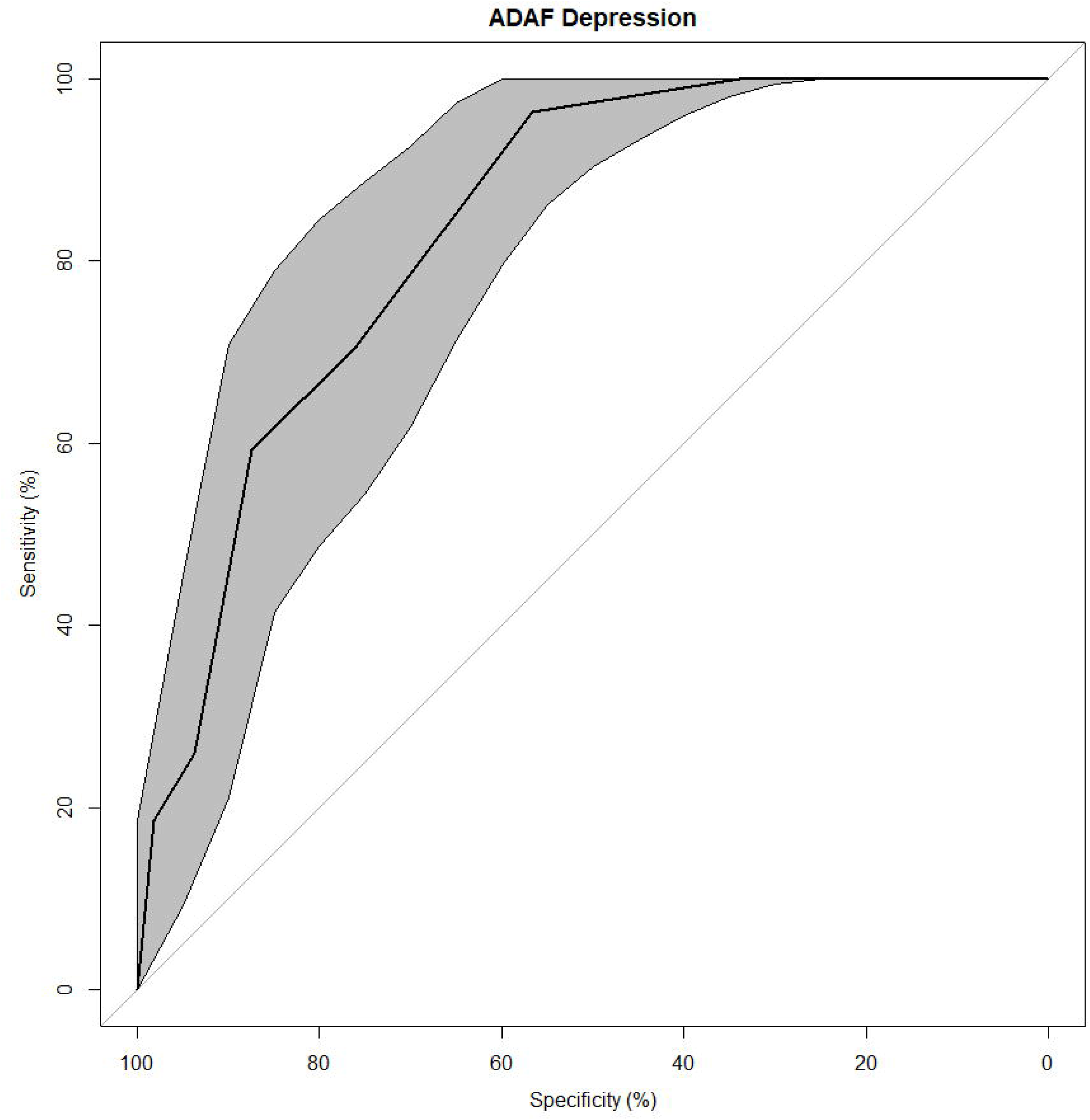
Receiver operating characteristic (ROC) curves for ADAF depression. Area under the ROC curve with 95% confidence interval (CI) is 0.84 (95% CI, 0.77-0.90).

**Figure 3.**
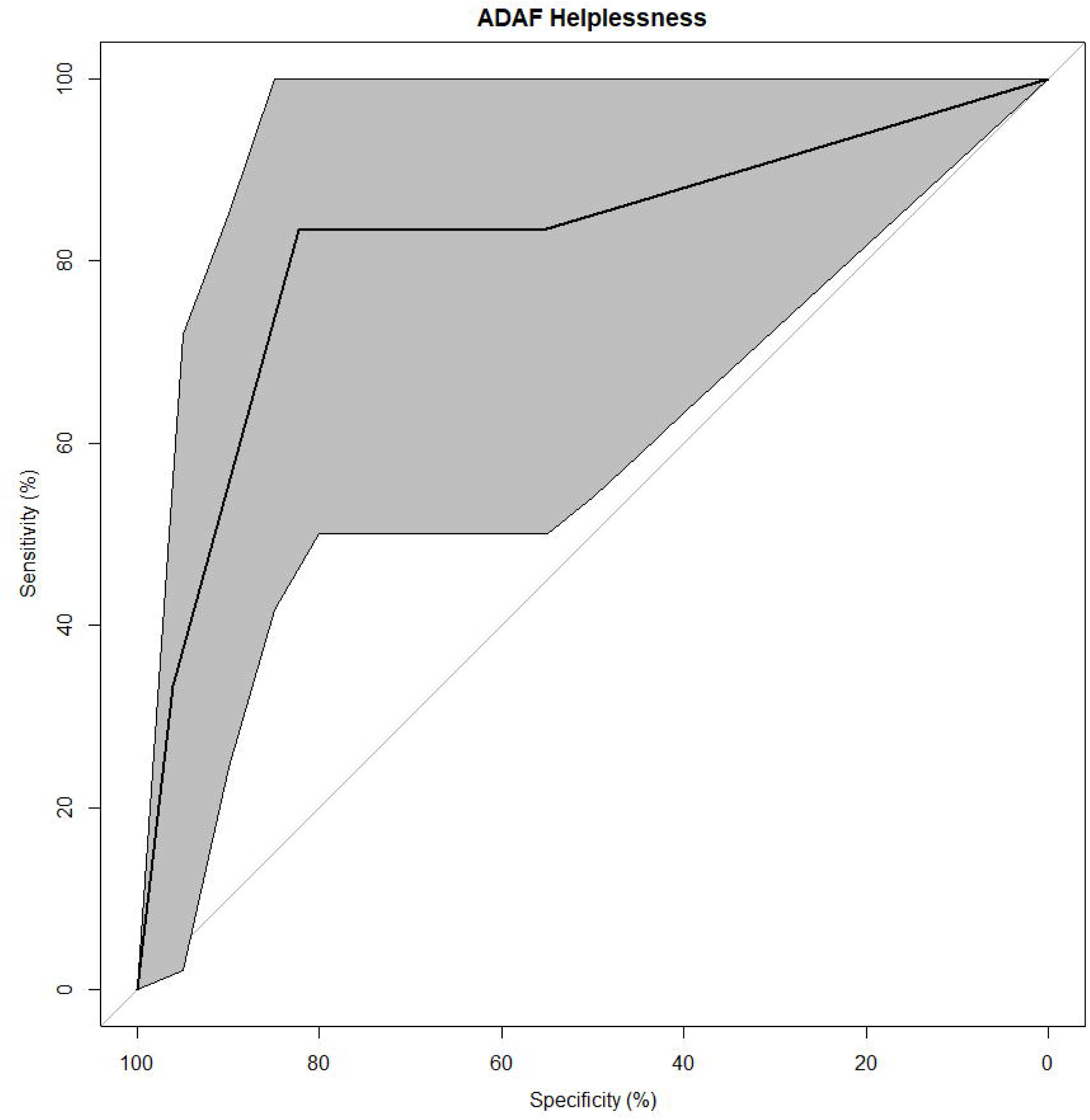
Receiver operating characteristic (ROC) curves for ADAF helplessness coping. Area under the ROC curve with 95% confidence interval (CI) is 0.82 (95% CI, 0.60-0.99).

**Figure 4.**
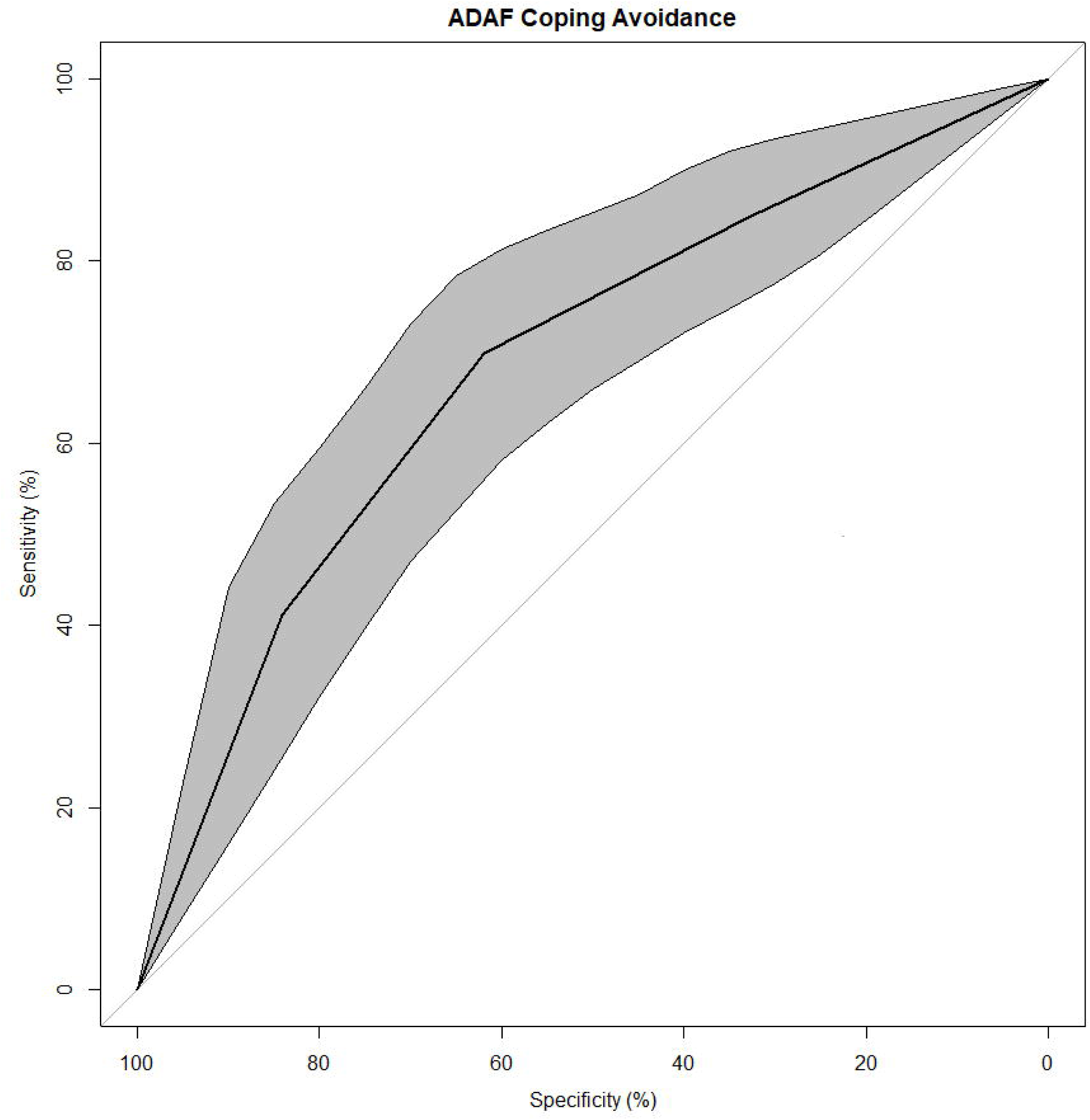
Receiver operating characteristic (ROC) curves for ADAF avoidance coping. Area under the ROC curve with 95% confidence interval (CI) is 0.68 (95% CI, 0.61-0.76).

According to these analyses, the ADAF for depression, in contrast to their gold standard taken as reference (HADS), presents a sensitivity that oscillates between 75% and 90.9%, and a specificity between 72.7% and 78.8%. According to these data, and to facilitate the establishment of the cut-off of depression measured with the ADAF, it is chosen for an overall score ≥3, showing a similar sensitivity (70.37%) and specificity (76.1%). Thus, it is determined that patients with scores ≥3 would present depressive symptoms.

Regarding anxiety, a cut-off of 2 was selected by the metric score and showing lesser sensitivity (67.2%) than specificity (89.6%). When coping avoidance scores is assessed (taking as gold standard reference the MiniMAC Coping Avoidance), suggested cut-off is 2 with similar sensitivity (69.86%) and specificity (61.95%). Regarding coping helplessness (taking as gold standard reference the MiniMAC Coping Helplessness), the best performance cut-off is 2, with similar sensitivity (83.33%) and specificity (82.2%) (Table 2).

**Table 2.**
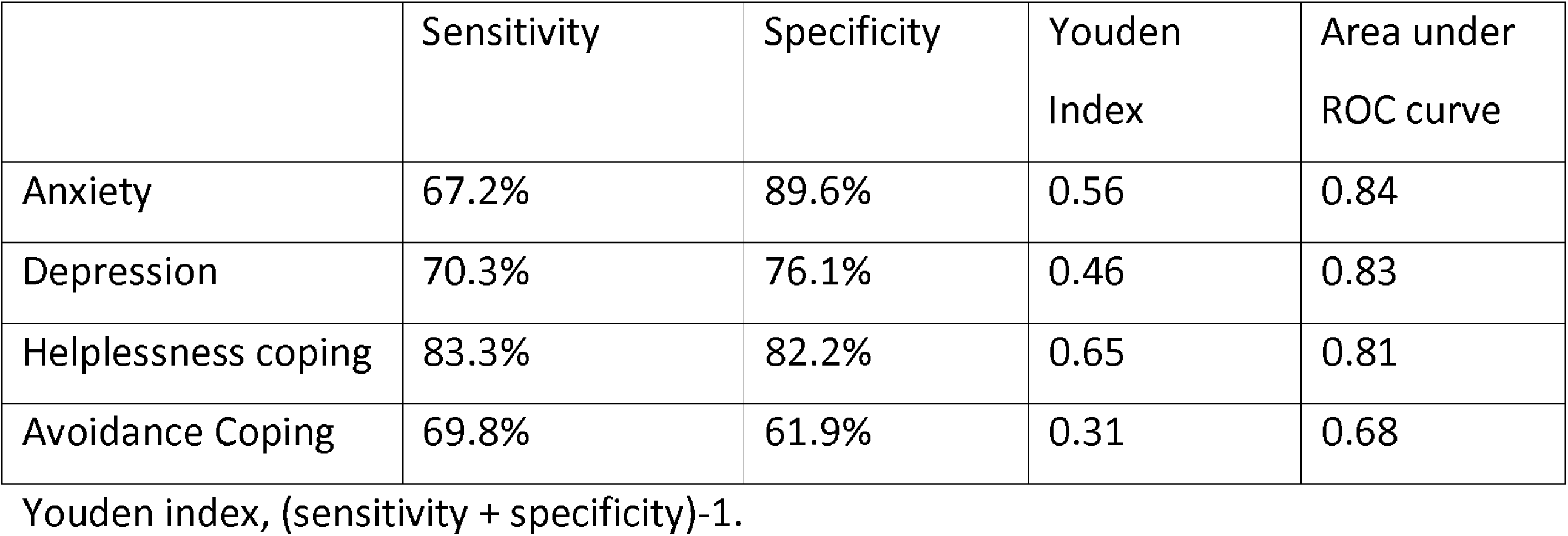
Operating Characteristics of the ADAF screening and gold-standars (HADS and MninMAC)

|  | Sensitivity | Specificity | Youden Index | Area under ROC curve |
| --- | --- | --- | --- | --- |
| Anxiety | 67.2% | 89.6% | 0.56 | 0.84 |
| Depression | 70.3% | 76.1% | 0.46 | 0.83 |
| Helplessness coping | 83.3% | 82.2% | 0.65 | 0.81 |
| Avoidance Coping | 69.8% | 61.9% | 0.31 | 0.68 |

Youden index, (sensitivity + specificity)-1.

## Discussion

In this multicenter study of patients with cancer, the ADAF demonstrated good diagnostic screening for emotional and coping problems in cancer, as illustrated by AUCs between 0.68 to 0.84. The systematic application of this rapid tool would allow patients with psychological problems to be easily identified in clinical practice, and therefore aid healthcare professionals to adjust their communication to the psychological state of the patient, as well as identify patients that call for specialised care.

The context of the study is the rising burden of oncological diseases. The experience of the disease in the patient is multidimensional. Patient-reported outcomes (PROs) are used to assess patient health across broad areas including symptoms, physical functioning, work and social activities, and mental well-being. In fact, the use of PROs and specifically related with the psychological state, which represent the needs and problems of the patients and can be obtained from our healthcare activity, would allow us to know general indicators of health outcomes, to analyse the evolution of a patient’s outcome, and furthermore to compare different and heterogeneous, clinical situations. In this sense, we think that ADAF is a simple and applicable screening tool in different oncological diseases.

If psychological needs are not addressed, regardless of when they arise, they then predict later stress, more anxiety and depressive symptoms, low quality of life, increased adverse effects, poor adherence and more physical symptoms [25,26]. It can be a first approach to referring patients in need for a more thorough evaluation by mental health professionals. Therefore, strategies for early identification should be a priority in oncology.

The ADAF results as a combination among two important dimensions in cancer, which are negative emotions (anxiety and depression) and coping. One of the advantages of this questionnaire is that unites the two dimensions in a single screen.

Maladaptive coping screening quickly identifies which patients may have significant difficulties in their process of coping with illness and adapting to treatments. Working on the helplessness coping style means enhancing the patient ability to control. Thus, people who believe that they can do something for their health will initiate more health behaviours, will become more involved in their rehabilitation and will be more relented to failures [27]. In general, they will cope more actively.

Lack of perceived control is associated with depression. It has been observed that depressed subjects have lower perception of control, being also associated with escape and avoidance [28]. Without perceived control, distress is increased, and active coping strategies are not carried out [29].

The use of this tool also allows professionals a better exploration of the psychological state and adjustment to the disease of their patients, to offer support and provide education and information to all patients and their families about psychological problems and its treatments, regardless of whether a need for referral to mental health. Furthermore, patients feel that their doctor is concerned not only to medical variables, but also to other aspects of their illness. This fact may have a favourable impact on patient’s psychological wellbeing and on their relationship with the health professional.

Some aspects of the ADAF potentially increase its acceptability compared with the most efficient tools, which is usually considered too lengthy for routine use. It has only five items in its full form and takes approximately 2 minutes to complete.

Another reason to choose short tools instead full scales is to avoid that people get tired and leave questionnaires incomplete, which is one of the risks of autocompleted measurements. By the end, the professional does not need to make a correction since they receive an immediate response based on the cut-off point. This response is automatic and informative, so when the patient scores above, it is clear that they need to be referred for psychological support, acting as a “red flag” in the higher scores. Moreover, recent evaluations have demonstrated reductions in healthcare utilization when institutions adhere to distress screening protocols [30].

Strengths of our study include the multicenter design with a large representative sample of patients from all treatment settings. The sample included patients with different diseases, stages of disease, receiving different medical treatments, and at different stages of the disease process, so the generalizability of findings is good. Nevertheless, our study also has some limitations, including it is a cross-sectional design and the fact that the two measures are self-reports. Future research should explore the temporal stability and the change sensitivity of the instrument.

Another limitation is that including different subtypes of patients and analysing them as a group may obviate the intrinsic differences between them. Furthermore, although the ADAF has proven its reliability as a screening tool, a clinical approach is necessary to diagnoses and managing emotional or adjustment disorders.

In closing, emotional problems or maladaptive coping strategies are frequent in cancer, so it is necessary to implement screening or detection tools appropriate to the healthcare reality, since the early and simple detection of patients at risk will allow the professional to adjust the communication and approach of the patient from that moment, being suitable for making clinical decisions, follow-up, or referral to mental health care.

## Data Availability

Data are available upon request

## Declarations

## Acknowledgments

This work has been supported by Fundación Mylan para la salud. The authors thank the patient’s participation.

## Contributions

Conceptualization: MR, PP and LL; data collection: FS, MB, ID, and JV; data analysis: LL and GC; writing—original draft preparation: LL, GC, MR; writing—review and editing: MAP, PP, IJ, SE, EH, HO and JLB; supervision: LL and MR.

## Ethics declarations

### Conflict of interest

All authors had financial support from Fundación Mylan para la salud for the submitted work; no financial relationships with any organizations that might have an interest in the submitted work in the previous three years; no other relationships or activities that could appear to have influenced the submitted work.

### Ethical approval

This study was approved by the Ethics and Clinical Research Committee of the Hospital Clínico San Carlos (CEIC nº19 / 265-E),and was conducted in accordance with the ethical standards of the Declaration of Helsinki and its later amendments.

### Informed consent

Local ethical committee approved the use of anonymized historic data for the study and waived informed consent from patients.

## APPENDIX 1 ADAF screening questionnaire

Read each sentence and check the answer that best matches how you felt during the past week (0. Almost never 1. Few times 2. Many times 3. Almost always)

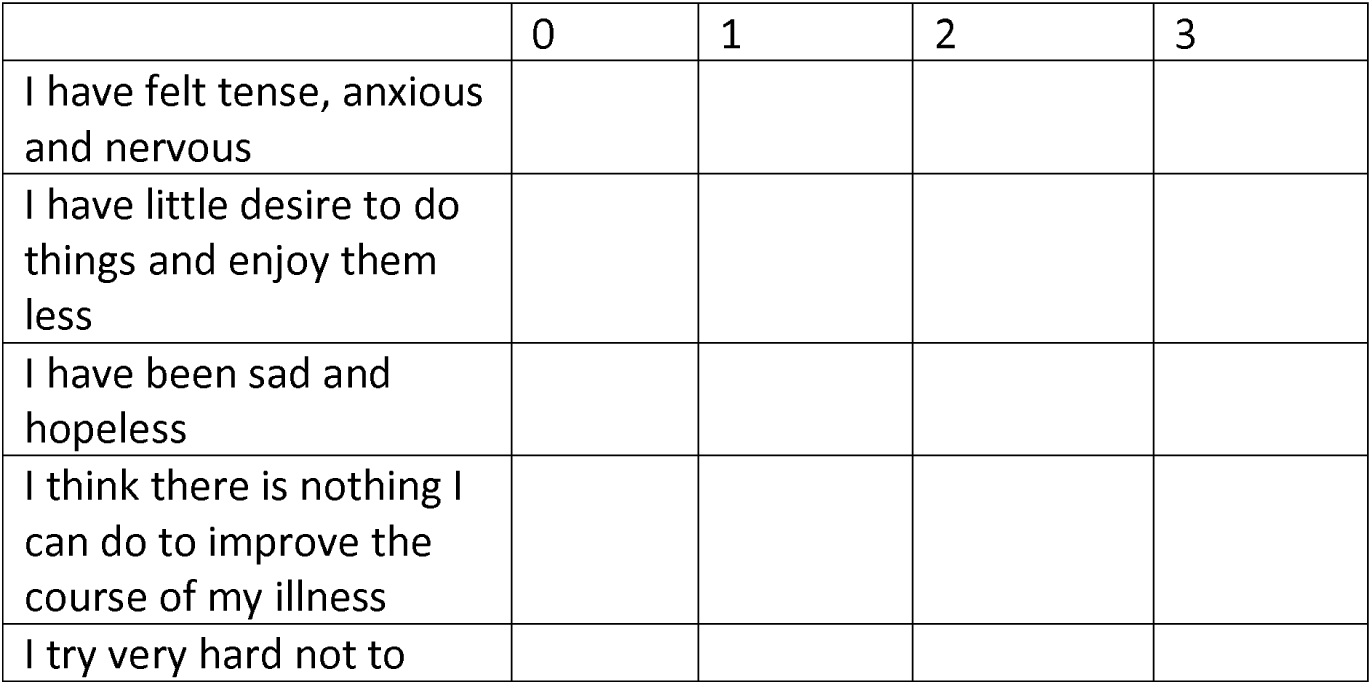
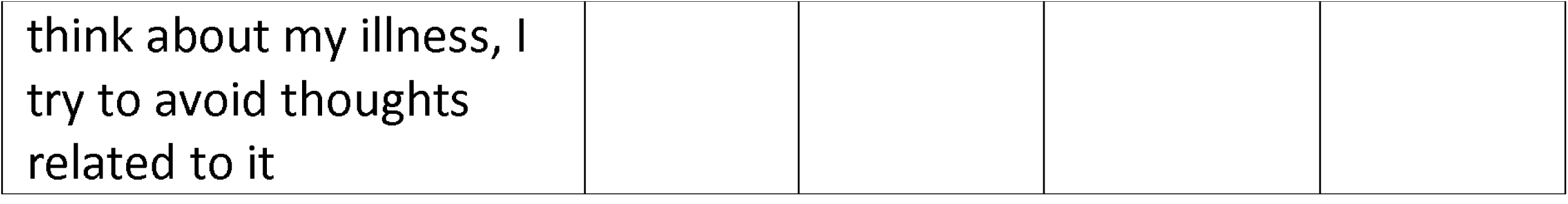

## Notes

### Competing Interest Statement

All authors had financial support from Fundacion Mylan para la salud for the submitted work; no financial relationships with any organizations that might have an interest in the submitted work in the previous three years; no other relationships or activities that could appear to have influenced the submitted work.

### Author Declarations

The research protocol was approved by the Ethics and Clinical Research Committee of the Hospital Clinico San Carlos (CEIC 19 / 265-E).

